# Second-line therapy following osimertinib in metastatic *EGFR*-mutated non-small cell lung cancer at an academic medical center

**DOI:** 10.1101/2025.10.29.25339070

**Authors:** Michael Rafizadeh, Stephanie Bogdan, Jonathan Lee, Christine Garcia, Ashish Saxena, Kathy Zhou, Bobak Parang

**Affiliations:** Division of Hematology and Oncology, Department of Medicine, Weill Cornell Medicine, New York, NY; Department of Population Health Sciences, Weill Cornell Medicine, New York, NY; Division of Medical Oncology, Department of Internal Medicine, The Ohio State University Comprehensive Cancer Center, Columbus, OH

**Author notes:** **Corresponding Author** Bobak Parang, MD, PhD, Assistant Professor, Division of Medical Oncology, Department of Internal Medicine, The Ohio State University Comprehensive Cancer, Center Biomedical Research Tower, 460 W 12^th^ Avenue Columbus, OH 43210.

**Keywords:** EGFR, non-small cell lung cancer, second line therapy, osimertinib, FLAURA2

## Abstract

**Purpose:** FLAURA2 demonstrated that adding chemotherapy to osimertinib improved overall survival compared with osimertinib monotherapy in metastatic *epidermal growth factor receptor*-mutated (*EGFR*-mut) non-small cell lung cancer (NSCLC). Notably, only 60% of patients in the osimertinib monotherapy arm received second-line therapy after discontinuing first-line osimertinib, raising the concern that FLAURA2 did not accurately reflect real-world practices at academic medical centers. We hypothesized that a higher proportion of patients on osimertinib monotherapy receive second-line therapy at academic medical centers in the United States (US).

**Patients and Methods:** This is a retrospective cohort study of 115 patients with metastatic *EGFR*-mut NSCLC treated with first-line osimertinib monotherapy at an academic medical center in the US from February 2018 to July 2024. Analyses included Kaplan-Meier survival estimation, the log-rank test, multivariate Cox regression, and the Kruskal-Wallis test.

**Results:** Most patients were female (74%) and had a history of never-smoking (69%). Fifty percent were Asian, and 93% of patients had adenocarcinoma histology. The median time to treatment failure (TTF) for all patients on first-line osimertinib was 25.3 months (95% CI: 18.6– 37.5). The median TTF was 16.3 months (CI: 13.3–22.0) for *TP53*-mutated patients and 42.3 months (CI: 36.9–NA) for *TP53* wild-type patients (log-rank test, P < 0.001). Of the 115 total patients, 66 (57.4%) discontinued first-line osimertinib. Of these 66 patients, 26 (39.4%) either died or pursued hospice. Forty (60.6%) of the 66 patients experienced progression of disease and subsequently received second-line therapy.

**Conclusions:** Only 61% of patients with metastatic *EGFR*-mut NSCLC received second-line therapy after osimertinib at our institution, confirming that the second-line therapy rates in the control arm of FLAURA2 are similar to practice patterns at our US academic medical center.

## Introduction

The FLAURA2 trial demonstrated that adding chemotherapy to osimertinib improved overall survival (OS) compared with osimertinib monotherapy in metastatic *epidermal growth factor receptor*-mutated (*EGFR*-mut) non-small cell lung cancer (NSCLC).^1,2^ A widespread criticism of the trial is that only 60% of patients in the osimertinib monotherapy arm received second-line therapy after discontinuing first-line osimertinib, raising the concern that the control arm in FLAURA2 did not accurately reflect real-world practices and had limited applicability to academic medical centers in the United States (US).

Several recent studies have shown that real-world, post-progression treatment practices vary across the world and at different institutions. An Italian study reported that only 55% of patients who were treated with osimertinib received second-line treatment.^3^ An analysis of 30 institutions across Japan revealed that approximately 42% of patients received second-line treatment after osimertinib.^4^ More recently, a large retrospective study of community practices across the US using the Flatiron Health database showed that approximately 44% of patients received second-line therapy.^5^ A key gap in the literature is whether the rates of post-progression therapy in the FLAURA2 control arm are consistent with practices at academic medical institutions in the US and whether FLAURA2 applies to academic medicine.

We hypothesized that a higher proportion of patients on first-line osimertinib monotherapy receive second-line therapy at academic medical institutions in the US. To test this hypothesis, we conducted a retrospective cohort study at our urban academic medical center. Our primary objective was to determine the proportion of patients on first-line osimertinib who receive second-line therapy. Our secondary objectives were to report survival data for this cohort and to investigate what clinicodemographic and genetic variables affected outcomes.

## Patients and Methods

### Data collection and statistical analysis

Clinical, demographic, genetic, and treatment data for the first three lines of therapy were abstracted from medical records. Data were censored at the time of last follow-up or the cutoff for data collection (January 3, 2025). Date of diagnosis was defined as the date of collection of a specimen that underwent pathologic analysis confirming *EGFR*-mut NSCLC. *EGFR*-mut NSCLC was defined as tumors harboring an *EGFR* exon 19 deletion or *EGFR* L858R mutation. All patients underwent next-generation sequencing (NGS)-based mutational profiling. Time to treatment failure (TTF) was defined as the duration of time from initiation of treatment to discontinuation of treatment. If a patient continued therapy after disease progression but received an additional agent, the end date for the original therapy was recorded as the initiation date of the new therapy. Descriptive statistics were calculated, and the Kruskal-Wallis test, Kaplan-Meier survival estimation, the log-rank test, and multivariate Cox regression were performed. Since *TP53* status was not known for all patients, hazard ratios for *TP53* were estimated from a model that included *TP53* status, whereas hazard ratios for all other covariates were estimated from a model that excluded *TP53* status to maximize sample size. All confidence intervals are 95% unless otherwise specified.

### Informed Consent

The Institutional Review Board (IRB) of Weill Cornell Medicine and New York-Presbyterian Hospital approved the retrospective study protocol. Patient informed consent was waived by the IRB.

## Results

### Study population

We identified 258 patients evaluated at our institution for metastatic *EGFR*-mut NSCLC from February 2018 to July 2024 through an institutional database. Patients were excluded if they were not treated with first-line osimertinib monotherapy (*n* = 55), were not treated at our institution (*n* = 29), did not harbor classical *EGFR* mutations (L858R or exon 19 deletion) (*n* = 19), were lost to follow-up before initiation of second-line therapy (*n* = 20), had a concurrent active malignancy (*n* = 8), discontinued first-line osimertinib due to intolerance of side effects (*n* = 6), had inadequate documentation (*n* = 4), had interstitial lung disease (*n* = 1), or had a duration of >3 months from diagnosis to initiation of osimertinib (*n* = 1) (**Figure 1**).

**Figure 1.**
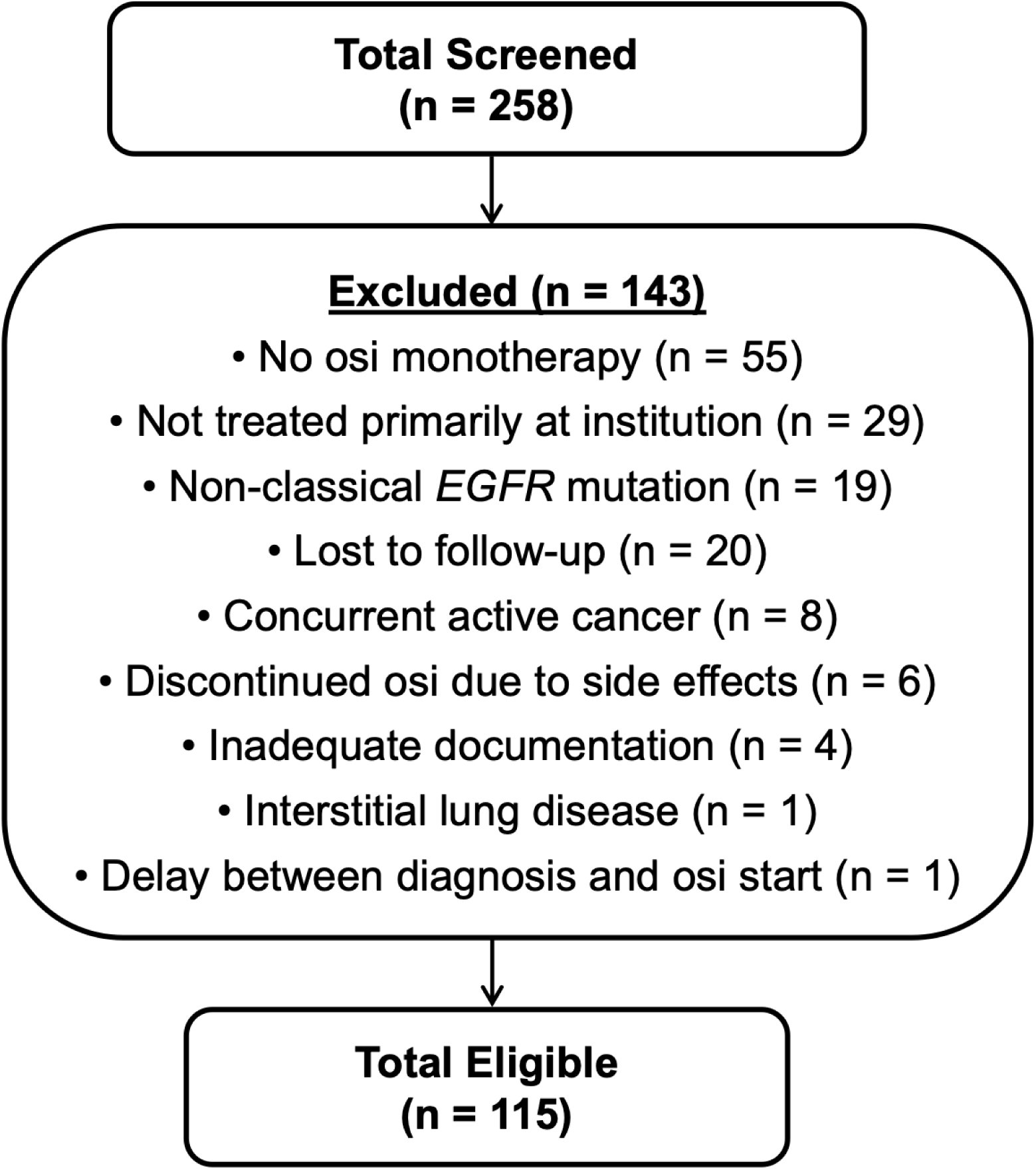
CONSORT flow chart detailing patient screening and exclusion

Of the 115 evaluable patients, the median age at diagnosis was 69 years. Most patients were female (*n* = 85, 74%), had a history of never-smoking (*n* = 79, 69%), and had adenocarcinoma histology (*n* = 107, 93%). Fifty percent (*n* = 57) of patients were Asian, 37% (*n* = 42) had central nervous system (CNS) involvement at baseline, 54% (*n* = 62) had an L858R mutation, 46% (*n* = 53) had an exon 19 deletion, and 54% (*n* = 62) had a concurrent *TP53* mutation at diagnosis. Demographic information is specified in greater detail in **Table 1**.

**Table 1.** Demographic, disease, and mutational data for the cohort.

| Characteristic | No. (%) |
| --- | --- |
| Median age at diagnosis – yr (range) | 69 (42–94) |
| Sex – no. (%) |  |
| Male | 30 (26) |
| Female | 85 (74) |
| Race |  |
| Asian | 57 (50) |
| White | 24 (21) |
| Other | 9 (8) |
| Black | 14 (12) |
| Declined to Answer | 11 (10) |
| Ethnicity |  |
| Non-Hispanic | 88 (77) |
| Hispanic | 13 (11) |
| Declined to Answer | 14 (12) |
| Smoking Status |  |
| Mild | 23 (20) |
| Moderate | 8 (7) |
| Heavy | 5 (4) |
| Never-smoker | 79 (69) |
| Histology |  |
| Adenocarcinoma | 107 (93) |
| Squamous cell carcinoma | 3 (3) |
| Adenosquamous | 1 (1) |
| Large cell carcinoma | 1 (1) |
| Neuroendocrine carcinoma | 1 (1) |
| NSCLC not otherwise specified | 2 (2) |
| CNS Disease at Diagnosis |  |
| Yes | 42 (37) |
| No | 73 (63) |
| EGFR Mutation |  |
| L858R | 62 (54) |
| Exon 19 deletion | 53 (46) |
| TP53 Mutation Status |  |
| TP53-Mutated | 62 (54) |
| TP53 Wild-type | 50 (43) |
| TP53 Unknown | 3 (3) |

### Survival outcomes

Median time to treatment failure (mTTF) for all patients on first-line osimertinib (*n* = 115) was 25.3 months (CI: 18.6–37.5). At the time of data cutoff, the event rate for discontinuation of first-line osimertinib was 57.4%. For patients who reached second-line (*n* = 40) and third-line therapy (*n* = 22), mTTF on second-line and third-line therapy was 4.7 months (CI: 3.4–7.1) and 4.8 months (CI: 2.7–7.1), respectively. Median OS (mOS) for all patients was 38.1 months (CI: 30.5–NA). At the time of data cutoff, the event rate for death was 44.3%, and the median follow-up time was 22.1 months (**Figure S1**).

Median age of *TP53*-mutated (*TP53*-mut) patients was 66 years, compared with 73 years for *TP53* wild-type patients. *TP53*-mut patients (*n* = 62) had a significantly lower mTTF than *TP53* wild-type (*n* = 50) patients (16.3 months [CI: 13.3–22.0] vs. 42.3 months [CI: 36.9–NA]; *P* < 0.001). Similarly, *TP53*-mut patients also had significantly lower mOS compared with *TP53* wild-type (27.2 months [CI: 22.2–42.4] vs. 63.5 months [CI: 42.4–NA]; *P* = 0.002) (**Figure S2**). Three patients had unknown *TP53* mutation status and were excluded from this analysis.

Multivariate Cox regression analysis showed that, after controlling for age, gender, race, presence of CNS disease at diagnosis, smoking status, type of *EGFR* mutation, and *TP53* mutation status, only age and *TP53* status remained independently significant for OS. For TTF, only *TP53* status remained independently significant. The adjusted hazard ratios (HR) for death and treatment failure for *TP53*-mut patients compared with *TP53* wild-type were 4.02 (CI: 2.01– 8.02, P < 0.001) and 4.99 (CI: 2.68–9.28, P < 0.001), respectively (**Figure S3**).

### Proportion of patients treated with second-line therapy after osimertinib

Of the 115 total patients, 57.4% (*n* = 66) discontinued first-line osimertinib. Of these 66 patients, 39.4% (*n* = 26) died or pursued hospice, while 60.6% (*n* = 40) experienced disease progression and received second-line therapy. Of the 115 total patients, 42.6% (*n* = 49) continued on first-line osimertinib at the time of data cutoff; among them, 20.0% (*n* = 23) continued the regimen for at least 24 months, while 22.6% (*n* = 26) had a treatment duration of less than 24 months.

### Age and ECOG status

Patients who did not receive second-line therapy had a median age (IQR) of 77.7 (70.5– 82.2), compared with 64.9 (60.3–71.3) for those who did receive second-line therapy and 69.2 (61.5–78.4) for patients continuing first-line osimertinib (Kruskal-Wallis test: *P* < 0.001).

Of the 68 patients with an ECOG performance status of 0-1 at baseline, 54.4% (*n* = 37) discontinued first-line osimertinib. Of these 37 patients, 62.2% (*n* = 27) received subsequent therapy, while 37.8% (*n* = 10) did not receive second-line therapy. Of the 27 patients with an ECOG performance status of 2+ at baseline, 70.4% (*n* = 19) discontinued first-line osimertinib. Of the 19 who discontinued first-line osimertinib, 9 (47.4%) received subsequent therapy, while 52.6% (*n* = 10) did not receive second-line therapy. ECOG status was not available for 17.4% (*n* = 20) of patients.

### TP53 mutation impact

We subsequently asked whether *TP53* mutations affect second-line therapy rates. We hypothesized that patients with a concurrent *TP53* mutation had lower rates of second-line therapy after osimertinib. Of the 62 patients who carried a *TP53* mutation, 71.0% (*n* = 44) discontinued first-line osimertinib. Of these patients, 29.5% (*n* = 13) died or pursued hospice, and 70.5% (*n* = 31) experienced disease progression and subsequently received second-line therapy. Among the 50 *TP53* wild-type patients, 40.0% (*n* = 20) discontinued first-line osimertinib. Of these 20 patients, 11 (55.0%) died or pursued hospice, and 9 (45.0%) had disease progression and subsequently received second-line therapy. Three patients had unknown *TP53* status (**Table 2**).

**Table 2.**
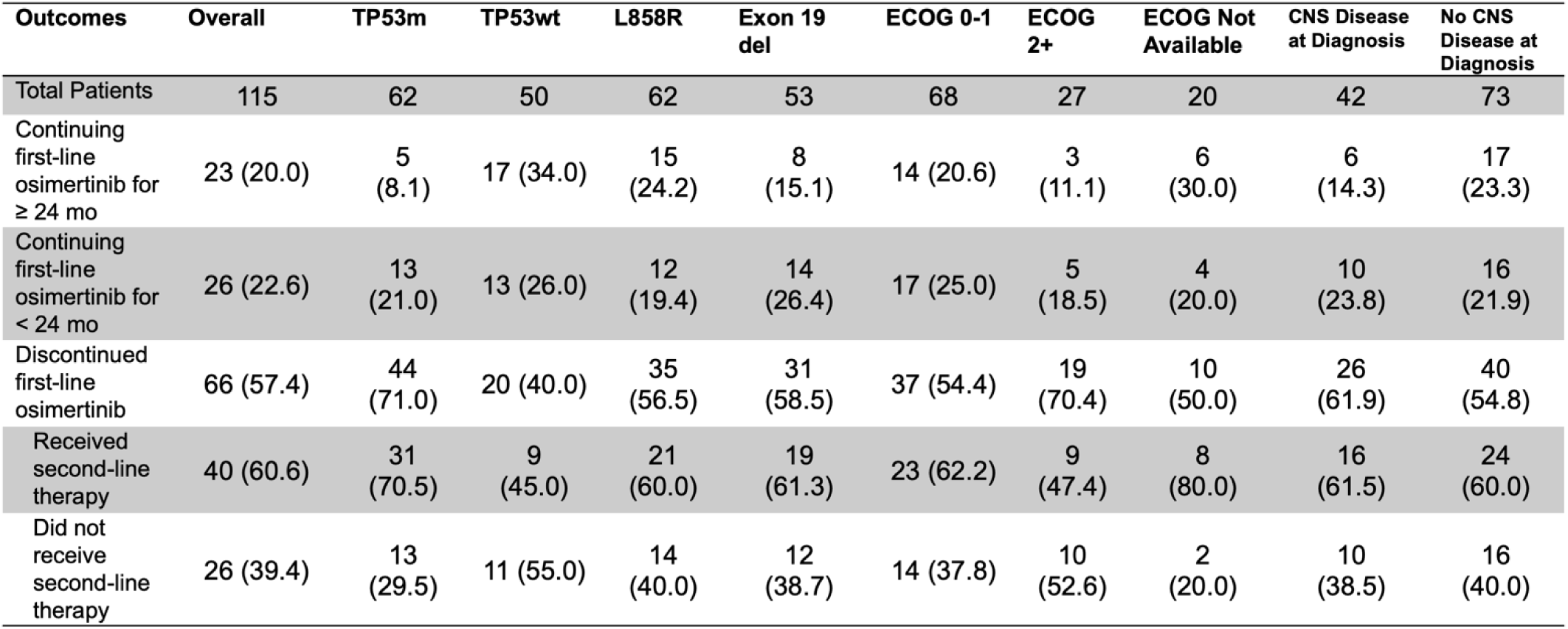
Outcomes after first-line therapy.

| Outcomes | Overall | TP53m | TP53wt | L858R | Exon 19 del | ECOG 0-1 | ECOG 2+ | ECOG Not Available | CNS Disease at Diagnosis | No CNS Disease at Diagnosis |
| --- | --- | --- | --- | --- | --- | --- | --- | --- | --- | --- |
| Total Patients | 115 | 62 | 50 | 62 | 53 | 68 | 27 | 20 | 42 | 73 |
| Continuing first-line osimertinib for $\geq 24$ mo | 23 (20.0) | 5 (8.1) | 17 (34.0) | 15 (24.2) | 8 (15.1) | 14 (20.6) | 3 (11.1) | 6 (30.0) | 6 (14.3) | 17 (23.3) |
| Continuing first-line osimertinib for $< 24$ mo | 26 (22.6) | 13 (21.0) | 13 (26.0) | 12 (19.4) | 14 (26.4) | 17 (25.0) | 5 (18.5) | 4 (20.0) | 10 (23.8) | 16 (21.9) |
| Discontinued first-line osimertinib | 66 (57.4) | 44 (71.0) | 20 (40.0) | 35 (56.5) | 31 (58.5) | 37 (54.4) | 19 (70.4) | 10 (50.0) | 26 (61.9) | 40 (54.8) |
| Received second-line therapy | 40 (60.6) | 31 (70.5) | 9 (45.0) | 21 (60.0) | 19 (61.3) | 23 (62.2) | 9 (47.4) | 8 (80.0) | 16 (61.5) | 24 (60.0) |
| Did not receive second-line therapy | 26 (39.4) | 13 (29.5) | 11 (55.0) | 14 (40.0) | 12 (38.7) | 14 (37.8) | 10 (52.6) | 2 (20.0) | 10 (38.5) | 16 (40.0) |

### Second- and third-line therapy after osimertinib

Second-line treatment regimens most commonly included chemotherapy (75%), continuation of osimertinib in addition to another agent (25.0%), immune checkpoint inhibitors (ICIs) (17.5%), vascular endothelial growth factor (VEGF) inhibitors (7.5%), and amivantamab (7.5%) (**Table 3**). Third-line treatment regimens most commonly included chemotherapy (50.0%), osimertinib with or without another agent (31.8%), ICIs (13.6%), and VEGFR2 inhibitors (13.6%). Since patients often received concurrent treatments, the sum of the percentages exceeds 100% (**Table 4**).

**Table 3.**
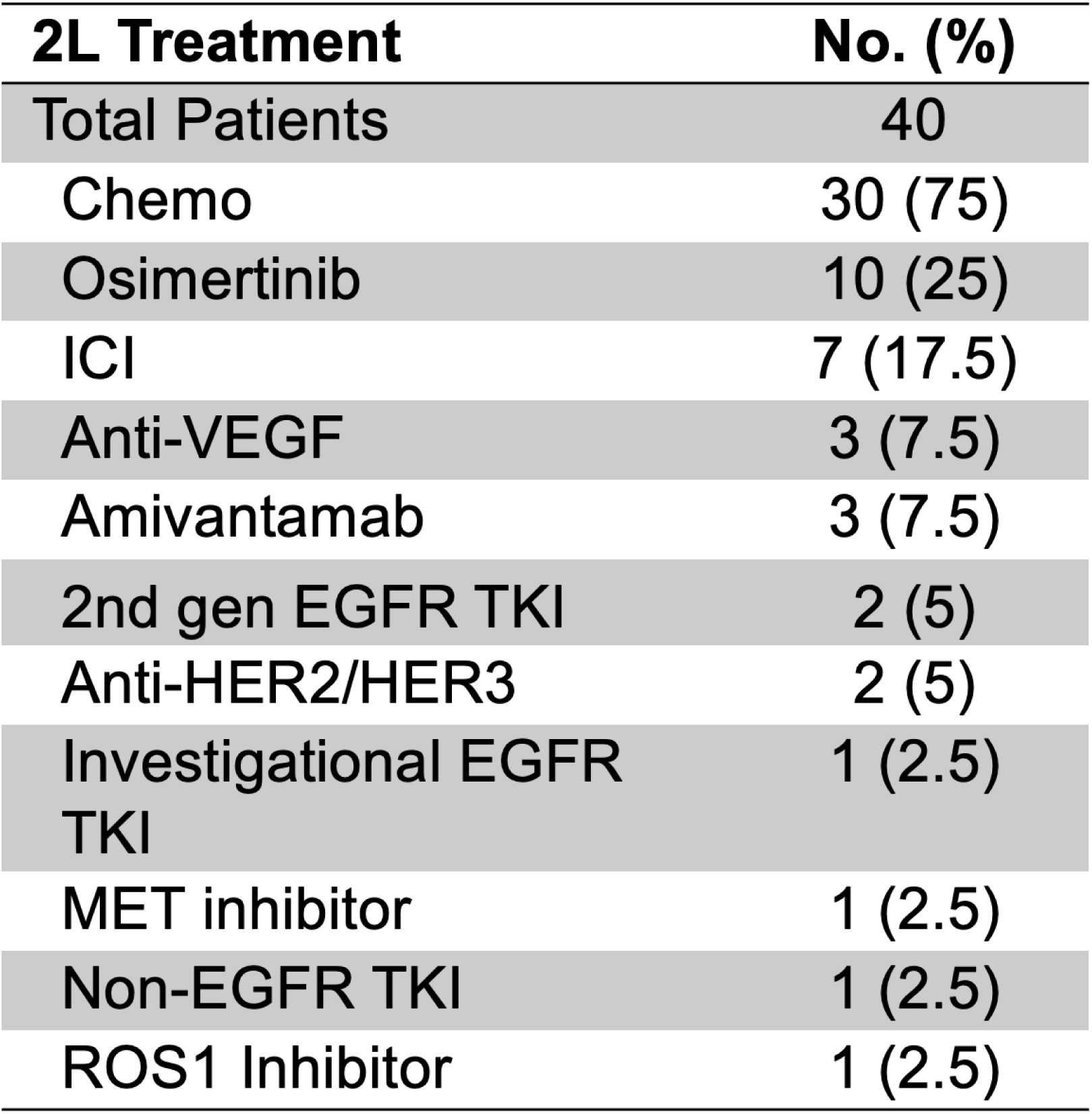
Second-line treatment regimens.

**Table 4.** Third-line treatment regimens.

| **3L Therapy** | **No. (%)** |
| --- | --- |
| Total Patients | 22 |
| Chemo | 11 (50) |
| Osimertinib | 7 (31.8) |
| Anti-VEGFR2 | 3 (13.7) |
| ICI | 3 (13.7) |
| Anti-TROP2 | 2 (9.1) |
| MET inhibitor | 1 (4.5) |
| MEK inhibitor | 1 (4.5) |
| BRAF inhibitor | 1 (4.5) |
| 2nd gen EGFR TKI | 1 (4.5) |
| Anti-HER3 | 1 (4.5) |
| Anti-VEGF | 1 (4.5) |
| Amivantamab | 1 (4.5) |

Of the 40 patients who received second-line therapy, 6 (15.0%) continued on second-line therapy at the time of data cutoff. One patient (2.5%) was lost to follow-up on second-line therapy. Thirty-three patients (82.5%) discontinued second-line therapy. Of these 33 patients, 9 (27.3%) died or pursued hospice, 2 (6.1%) discontinued treatment due to side effects, 1 (3.0%) stopped treatment due to withdrawal from a clinical trial, and 21 (63.6%) experienced disease progression. Two patients (6.1%) who progressed died shortly after progression and did not receive third-line therapy. Altogether, 22 patients (66.7%) discontinued second-line therapy and received third-line therapy.

Of the 22 patients who received third-line therapy, 2 (9.1%) continued on third-line therapy at the time of data cutoff. One patient (4.5%) was lost to follow-up on third-line therapy. Nineteen patients (86.4%) discontinued third-line therapy. Of these 19 patients, 4 (21.0%) died or pursued hospice, 1 (5.3%) discontinued treatment due to side effects, and 14 (73.7%) experienced disease progression (**Figure 2**).

**Figure 2.**
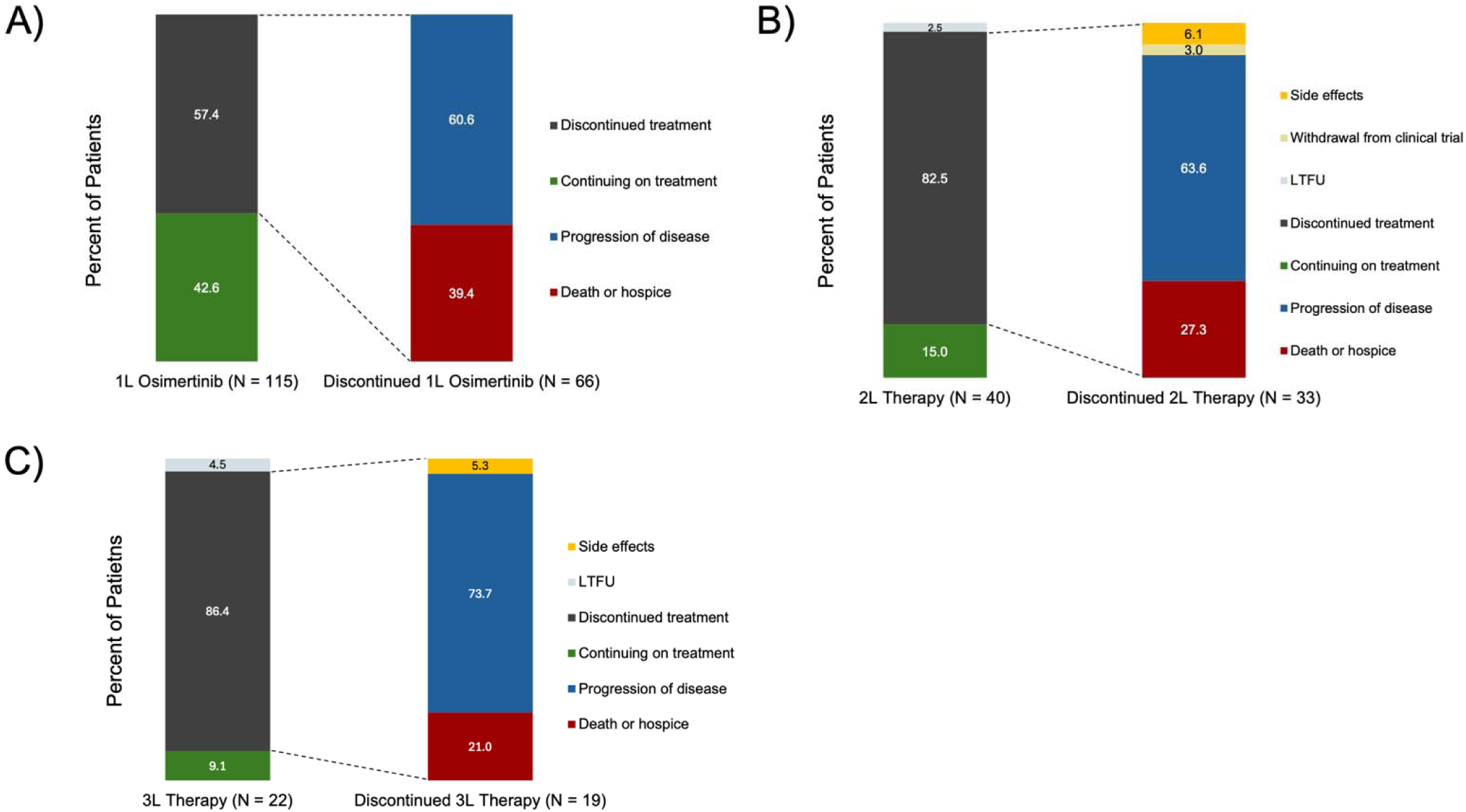
Outcomes following the first three lines of therapy. Patient outcomes after (a) discontinuation of first-line osimertinib (b) discontinuation of second-line therapy and (c) third line-therapy. LTFU=Lost to follow up.

## Discussion

The FLAURA2 trial ignited a debate about whether the control arm received standard of care according to respective geographical practices. International studies and retrospective reports of community practices in the US have shown similar rates of subsequent therapy after progression on osimertinib therapy. Whether FLAURA2 is similar to academic practices was not clear. To our knowledge, this is the first report showing academic practice patterns of second-line therapy after osimertinib. To our surprise, our study showed that only 61% of patients with metastatic *EGFR*-mut NSCLC received second-line therapy after osimertinib at our academic medical center. Our results show that second-line therapy rates in the control arm of FLAURA2 are similar to practice patterns at our US academic medical center as well as those reported in other studies.^6–9^

Unsurprisingly, the median age of those who did not receive second-line therapy was older than those who did, and a lower percentage of patients with ECOG performance status of 2+ received second-line therapy. Our study cohort was diverse and, compared with the cohort of FLAURA2, had an older median age at diagnosis by 8 years, roughly 10% fewer Asian patients, roughly 10% more women, and a similar proportion of patients with CNS involvement at baseline.

Consistent with prior literature, our study also shows that *TP53*-mut patients have inferior TTF and OS compared with *TP53* wild-type patients treated with first-line osimertinib.^6, 10^ Surprisingly, compared with *TP53* wild-type patients, more *TP53*-mut patients received second-line therapy (71% vs. 45%). We theorize that this may be due to the older baseline age of our *TP53* wild-type cohort or the nature of the disease progression; *TP53*-mut patients experience progression earlier on when they are better able to tolerate additional treatment, whereas *TP53* wild-type patients progress later when they are frailer. This observation must also be qualified by the limited sample size.

Uptake of second-line therapy depends on multiple factors, including patient preference, tumor biology, and the efficacy of first-line therapy. Despite the significant breakthroughs in lung cancer over the last 2 decades, studies across different driver mutations and treatment regimens show that approximately 50% of patients with metastatic NSCLC do not receive second-line therapy.^7–9, 11, 12^ Our work underscores the alarming rate of attrition between first-and second-line treatment in metastatic *EGFR*-mut NSCLC and highlights the minimal difference in community and academic clinical practices in achieving second-line therapy.

## Limitations

This was a real-world study primarily investigating the proportion of patients who receive second-line therapy after first-line osimertinib. The retrospective nature of the study carries inherent limitations. Additionally, this was a single-institution study that may or may not accurately reflect the findings of other institutions. We used TTF instead of progression-free survival (PFS) because progression events were not assessed formally by Response Evaluation Criteria in Solid Tumors (RECIST).^13^ We did not subclassify *TP53* mutations. Despite this, we observed a significant signal with respect to mTTF and mOS; homogenizing this group would expectedly bias any correlations to the null given the heterogeneity of prognostic significance of certain *TP53* mutations. Finally, genetic data after diagnosis or resistance patterns were not reliably available for our cohort.

## Data Availability

All data produced in the present work are contained in the manuscript

## Acknowledgments

The authors would like to thank Angela Dahlberg, editor in the OSUCCC Division of Medical Oncology, for editing this manuscript.

---

**Figure S1.**
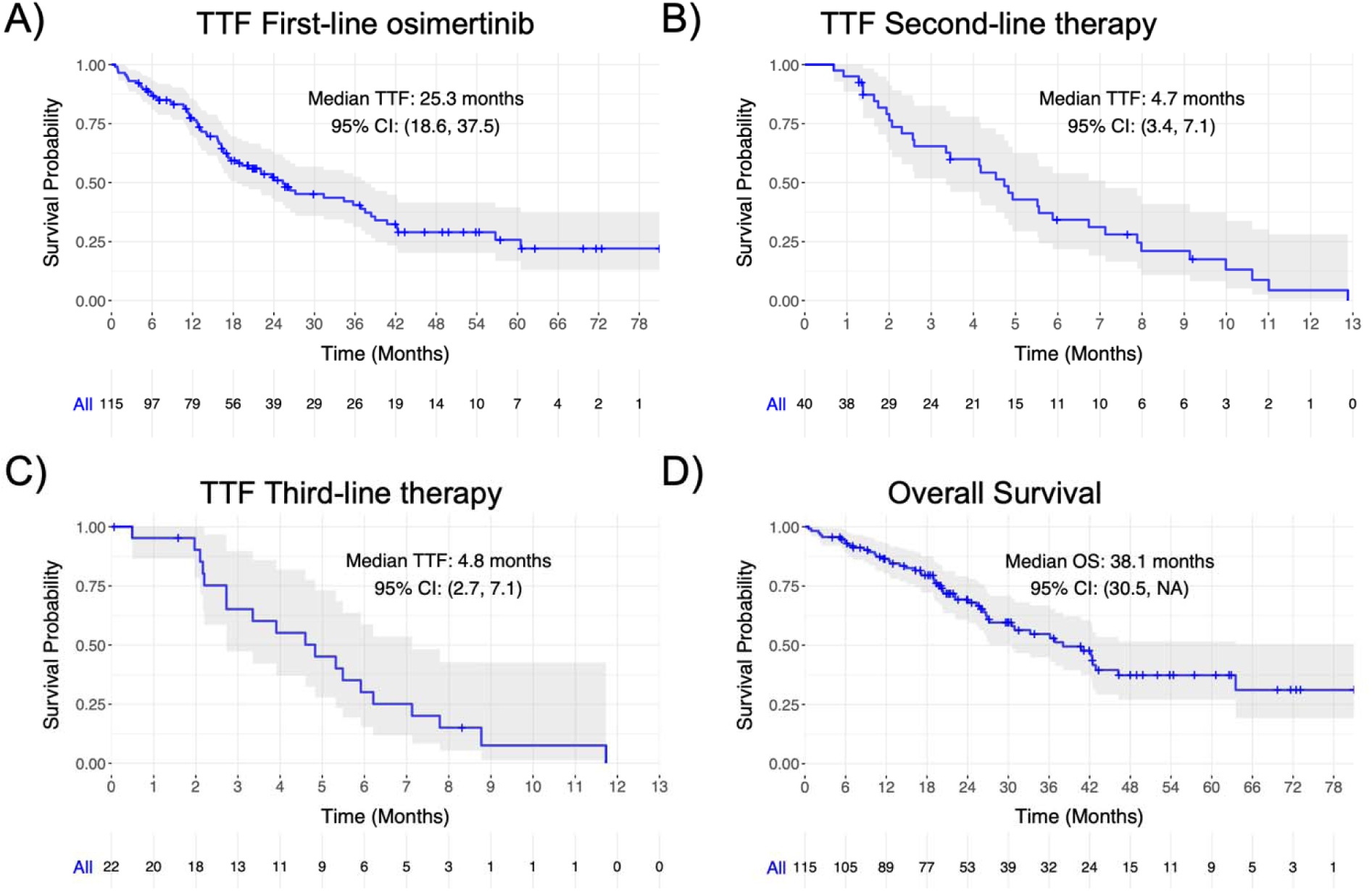
Kaplan-Meier survival analysis for time to treatment failure for (a) first, (b) second, and (c) third line therapy and (d) overall survival for all patients.

**Figure S2.**
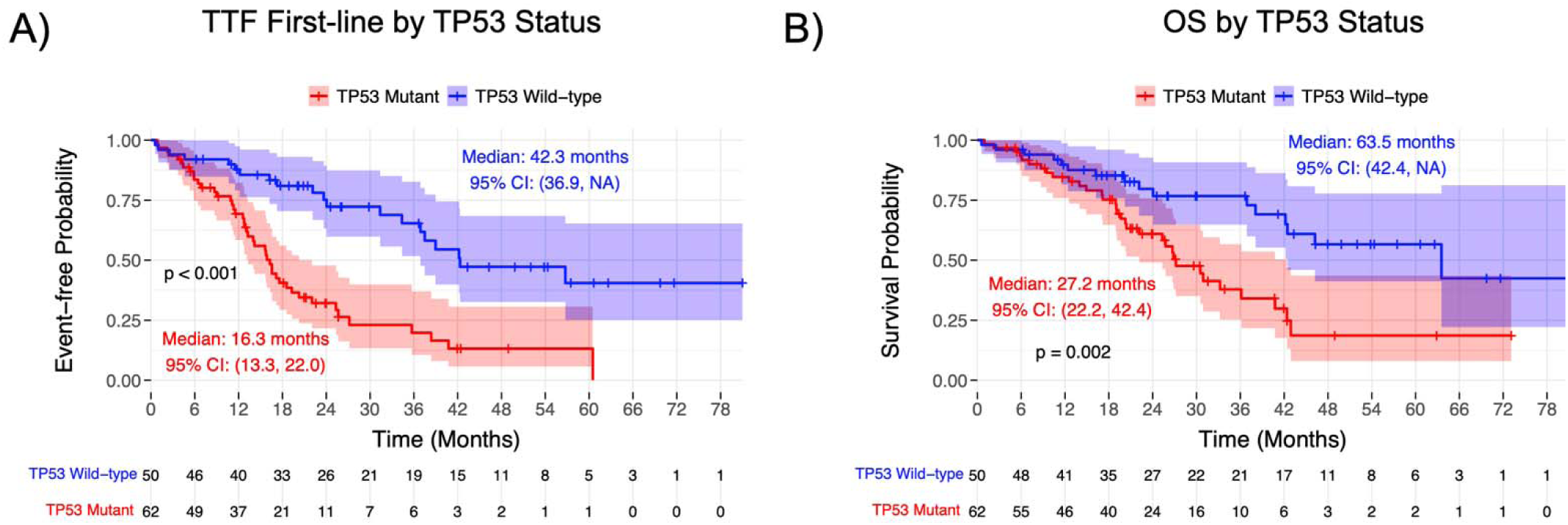
Kaplan-Meier survival analysis for (a) time to treatment failure and (b) overall survival (b) stratified by TP53 status.

**Figure S3.**
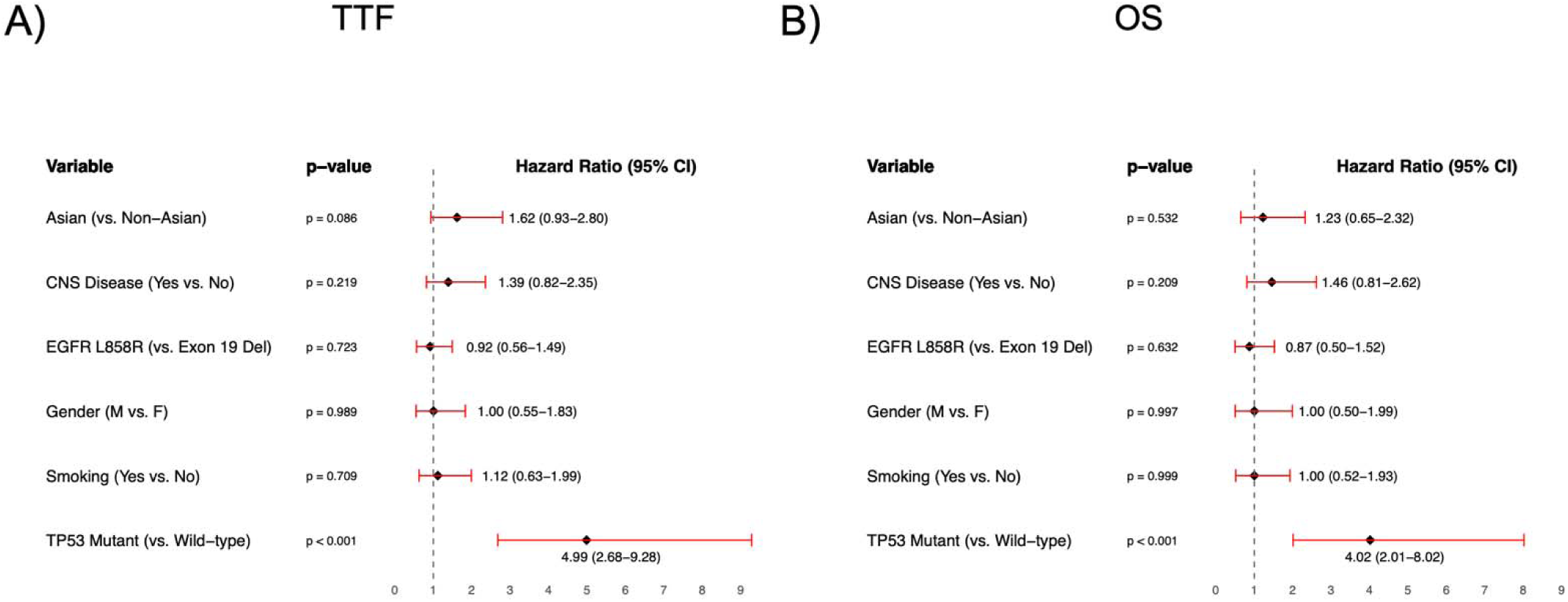
Forest plot for multivariate Cox regression analyses for time to treatment failure (a) and overall survival (b). Age was included in the analysis but is not shown graphically

